# Psilocybin Modulates TPJ Effective Connectivity during Out-of-Body Experiences

**DOI:** 10.1101/2025.06.24.25330245

**Authors:** Devon Stoliker, Fosco Bernasconi, Olaf Blanke, Adeel Razi

**Affiliations:** Turner Institute for Brain and Mental Health, Monash University, Clayton, VIC; Monash Biomedical Imaging, Monash University, Clayton, VIC; Laboratory of Cognitive Neuroscience, Neuro X Institute, Faculty of Life Sciences, Swiss Federal Institute of Technology (EPFL), Geneva, Switzerland; Department of Clinical Neurosciences, Geneva University Medical Center & Faculty of Medicine, University of Geneva, Geneva, Switzerland; Wellcome Centre for Human Neuroimaging, UCL, London, United Kingdom; CIFAR Azrieli Global Scholars Program, CIFAR, Toronto, Canada

**Keywords:** bodily sense of self, multisensory organisation, disembodiment, psychedelics, psilocybin, out of body experiences, OBE, bodily self-consciousness, effective connectivity, DCM, temporoparietal junction, temporal-parietal junction, TPJ, insula

## Abstract

Serotonergic psychedelics alter self-boundaries and can induce out-of-body experiences (OBEs)—the sense of being located outside one’s physical body. While OBEs also occur in clinical conditions and can be experimentally induced, their neural basis under psychedelics remains underexplored.

In an open-label, baseline-controlled MRI study of 62 healthy adults administered psilocybin, we examined effective connectivity changes in regions implicated in clinical and induced OBEs. Spectral dynamic causal modelling (spDCM) was applied to resting-state and music-listening scans to estimate connectivity changes from baseline and assess their consistency across contexts. Participants were grouped by self-reported OBE symptom intensity at the end of the dosing day.

In those reporting high-intensity OBEs, psilocybin reduced effective connectivity from the right to left anterior insula and between the right anterior insula and right temporoparietal junction (TPJ), inhibiting these connections across both scan types. These changes parallel known disruptions in TPJ–insula circuits linked to OBEs in clinical and experimental settings, particularly in the right hemisphere. Our findings highlight how psilocybin-induced disembodiment corresponds to altered effective connectivity and demonstrate the utility of spDCM for mapping causal neural dynamics underlying bodily self-consciousness.

## Introduction

### Understanding the effects of context

The subjective experience of inhabiting a body is a cornerstone of perceptual awareness. This bodily awareness is essential for interacting with the world and orienting oneself in physical space. However, what we consider foundational aspects of physical reality are underpinned by complex interplay of neuronal connections that shape our sense of body ownership and self-location. Alterations in these connections can profoundly affect the sense of body ownership, self-location, and first-person perspective (Blanke, Slater, & Serino, 2015), leading for example to so-called out-of-body experiences (OBEs) (Ionta et al., 2011).

OBEs belong to a category of autoscopic phenomena, alongside other autoscopic conditions including autoscopy, where one experiences a visual doppelgänger (Blanke, Landis, Spinelli, & Seeck, 2004; Brugger, 2002); and heautoscopy, which involves the doppelgänger as well as ambiguity whether the self is located at one’s physical body or the one of the Doppelgänger (Brugger, 2002). OBEs are distinct and involve a full experience of disembodiment. They are experienced as a separation of the self from the physical body, accompanied by a disintegration of the usual unity between the self, body, and one’s visual first-person perspective of the world (Metzinger, 2009). These experiences can include sensations of floating, detachment, and an external perspective, often dominated by an internally generated visual field (Winkelman, 2017). Additionally, OBEs can feature profound identity dissociations and are sometimes associated with near-death experiences (Blanke, Faivre, & Dieguez, 2016).

OBEs can be caused by a variety of conditions including transient psychological factors, sensory stimuli, and notably enduring neurological conditions such as epilepsy, migraines, and brain injuries (Blanke, 2004; Shushan, 2018). Interdisciplinary research, including neurological explorations of the clinical manifestations have provided a foundation for experimental inductions of OBEs. For instance, -irtual reality and robotic systems have been used to recreate out-of-body experiences, allowing their neurobiological underpinnings to be studied non-invasively in a controlled environment (Bernasconi et al., 2021; Blanke, 2012; Wu et al., 2024). Psilocybin, a common psychedelic molecule found in select mushroom species, offers a similar opportunity. Agonism of serotonergic receptors by the active metabolite of psilocybin (psilocin) can induce OBEs (Harduf, Panishev, Harel, Stern, & Salomon, 2023), making it and other psychedelics a valuable and novel tool for studying the neurobiological underpinnings of disembodiment phenomena.

A substantial body of research exploring OBEs highlights the function of specific brain regions localised to the temporoparietal junction (TPJ). The TPJ plays a crucial role in integrating multisensory information from the visual, somatosensory, proprioceptive, and vestibular systems, which is considered essential for creating a stable body representation and a sense of self being situated inside of the body (Blanke et al., 2016). Relatedly, the TPJ integrates multisensory signals to maintain bodily self-consciousness, including the experience of self-location, body position and perspective. Disruptions here can lead to out-of-body experiences, highlighting the TPJ’s role in separating self from environment (Dary et al., 2023; Olaf et al., 2005). Recent studies exploring the TPJ’s functionality in matching or mismatching expected and actual sensory inputs, a process crucial for maintaining the congruence of bodily self-consciousness, support this view (Doricchi, Lasaponara, Pazzaglia, & Silvetti, 2022). Disruptions in sensory integration at the TPJ, particularly in the right TPJ, as evidenced by studies from Blanke et al. (2004), (Ionta, Martuzzi, Salomon, & Blanke, 2014) and Ionta et al. (2011), are commonly observed in individuals experiencing varieties of autoscopic phenomenon, including OBEs. These studies suggests a critical role of the TPJ in modulating the experiences of self-location, first-person perspective and bodily awareness more generally, which are experiences that can be altered during psychedelic-induced OBEs (Studerus, Gamma, & Vollenweider, 2010). Disembodiment may depend on a disruptions to multisensory organisation of TPJ connectivity that underwrites the sensation of being situated inside or outside one’s body (Blanke & Arzy, 2005; Decety & Lamm, 2007; Lombardo et al., 2010). The function of the TPJ to integrate and segregate interoceptive and exteroceptive signals indicates it as a candidate for the dissolution of subject and object boundary commonly reported as in psychedelic experiences (H. D. Park & Blanke, 2019; Stoliker, Egan, Friston, & Razi, 2022).

TPJ anatomy extends from the end of the Sylvian fissure to the rostrolateral edge of the occipital cortex and is bordered by the posterior temporal lobe and the inferior parietal lobe, regions pivotal for integrating a myriad of sensory and cognitive inputs (Bukowski & Lamm, 2017). While TPJ lacks clear demarcation lines it can be divided into subregions based on their structural and functional connectivity, cytoarchitecture, or functional specialisations, and in general, encompasses several key areas: the supramarginal gyrus (SMG) located anteriorly, the superior temporal gyrus (STG) and sulcus (STS) below the Sylvian fissure, which are central to the TPJ and here referred to as the TPJ (Carter & Huettel, 2013). The angular gyrus (AG) is positioned posteriorly above, with its primary anatomical link to the temporoparietal junction (TPJ) via the superior longitudinal fasciculus (Caspers et al., 2011). The anterior portions of the insula connect to the TPJ through the uncinate fasciculus (Nieuwenhuys, 2012), enabling integration across interoceptive and multisensory networks.

The AG integrates cognitive and sensory inputs to generate a coherent body representation in space (Blanke, 2002), supporting body ownership and the ability to distinguish self-generated actions from external events (Tsakiris, 2005; Blanke, 2012). Disruptions in AG function—whether from pathology, trauma, electrical stimulation, or psychoactive substances—can impair sensory integration (Blanke, 2004). Direct evidence from electrical stimulation studies implicates the TPJ in OBEs, with stimulation inducing the experience of seeing oneself from an external perspective (Blanke et al., 2002). Further studies reveal that gaze direction and eye closure modulate these effects, linking the TPJ to visuo-spatial processing (Nakul, 2017). Complementary high-definition transcranial direct current stimulation (HD-tDCS) targeting the right AG alters self-location and perspective-taking (de Boer et al., 2020), highlighting AG connectivity as a contributor to the TPJ’s role in bodily self-consciousness.

The insula is similarly cited in OBE literature for its role integrating interoceptive states with external sensory inputs (H.-D. Park et al., 2018). It combines internal bodily signals with emotional information (Craig, 2009; Dary et al., 2023; Heydrich et al., 2018; H.-D. Park et al., 2018), and its functional coupling with the TPJ supports a unified sense of self in the environment (Dary et al., 2023; Ionta et al., 2014). Investigating connectivity within the TPJ-insula-AG network is essential, as each region uniquely contributes to the multisensory processes underpinning bodily self-consciousness and its disruption during OBEs.

Research linking psychedelics and OBEs is nascent. However, brain imaging studies indicate that psychedelics broadly affect connectivity in TPJ regions. For example, research shows that the administration of the synthetic psychedelic lysergic diethylamide acid (LSD) significantly increases the functional connectivity in the TPJ, AG, and insula (Tagliazucchi et al., 2016). This enhancement was suggested to synchronise activity across these regions, with the interpretation that this may potentially blur ordinarily experienced sensory distinctions. This could be indicative of the integrative role these areas play in modulating consciousness and bodily self-consciousness in particular. However, it is challenging to validate a link between subjective experiences and functional connectivity changes in the TPJ, as results can vary significantly based on the methods and scope of analysis (Preller et al., 2020; Smigielski, Scheidegger, Kometer, & Vollenweider, 2019). Moreover, much like experimental research that uses stimuli and signals to influence bodily self-perception, contextual factors under psychedelics, such as music, can modulate both subjective experience and corresponding connectivity (M. Kaelen et al., 2015; Mendel Kaelen et al., 2016; Stoliker et al., 2025). Context can therefore serve as a potential amplifier, point of reference and measure of consistency for psychedelic-induced OBEs.

Utilising a model of regions functionally connected with the TPJ that are implicated in both clinical and experimentally induced OBEs (see methods), we apply spectral dynamic causal modeling (spDCM) (Razi, Kahan, Rees, & Friston, 2015) to address the question of commonality between TPJ connectivity changes between psychedelics and previously reported non-pharmacological TPJ disruptions that induce OBEs. spDCM enables us to measure the directed neuronal connections in the TPJ under the influence of psychedelics during disembodiment, allowing the measurement of changes within excitatory-inhibitory balance of the TPJ that likely contribute to experiences of disembodiment, assessed by scales which measure self-reported feelings of disembodiment defined as floating, inhabiting space outside of one’s body, and the disappearance of one’s physical self (Studerus et al., 2010). Building on literature identifying the TPJs association with OBEs, our hypothesis posits that disruptions in the connectivity of the right TPJ play a crucial role in altering bodily awareness. Specifically, we expect that inhibition of the right TPJ effective connectivity is a key mechanism driving the subjective features of disembodiment during psychedelic experiences. To investigate the hypothesis that the bidirectional effective connectivity between the right TPJ and both the anterior insular (AI) and AG is altered during the subjective experience of psilocybin-induced disembodiment, we analysed data from resting state imaging sessions, followed by imaging sessions conducted with music. Performing a dual-condition analysis enabled us to evaluate the consistency of the observed changes in effective connectivity and to determine whether these changes could be influenced by auditory (music) stimuli. This research aims to deepen our understanding of the connectivity dynamics that underlie these profound experiences, making it a key area of interest in studies exploring the neural basis of consciousness and self-awareness. Moreover, characterising TPJ connectivity under psychedelics is essential for understanding the neurological bases of OBEs and has implications for treating perceptual and psychological disorders linked to these brain regions.

## Methods

### Participants

The data analysed in this study were collected as part of the PsiConnect (Psilocybin, Connectivity and Context) trial (Novelli et al., 2025; Stoliker et al., 2025), registered at anzctr.org.au (ACTRN12621001375842)) and was approved by the Monash University Human Research Ethics Committee (MUHREC). Sixty-five psychedelic naive healthy participants, aged 18-55, were recruited through online advertisements in Melbourne, Australia. All participants were deemed healthy after screening, which included medical history review, drug screening and physical examination. See Supplementary for more details.

### Design

An open label baseline-controlled study was performed. Testing days occurred approximately two weeks apart, and participants were orally administered 19mg psilocybin on the second testing day. The fixed dosage was selected based on previous findings that demonstrated no significant associations between the subjective effects of psilocybin and demographic variables such as body weight or sex (Garcia-Romeu, Barrett, Carbonaro, Johnson, & Griffiths, 2021). Approximately 80 minutes after oral administration of 19mg psilocybin, participants underwent a resting state scan (8 minutes, 505 volumes), followed by a music stimuli scan 7 minutes later, which featured an ethereal soundscape (11:24 minutes, 728 volumes).

Although the scan order was not randomised, variability in psilocybin onset is expected to account for any temporal effects related to OBE subjective experience. These sequences were selected for comparative analysis, to assess both consistency of the findings and robustness. Music scans were also selected based on participant reports that the music facilitated the subjective effects. Participants were asked to close their eyes during both scans. See Supplementary for details.

### MRI data acquisition and preprocessing

MRI data were acquired on a Siemens Skyra 3T whole-body scanner. A 32-channel receive head coil, and MultiTransmit parallel radio frequency transmission was used. Images were acquired using a whole-brain multi-echo multi-band sequence (TR of 910 ms, multi-echo TE of 12.60 ms, 29.23 ms, 45.86 ms, 62.49 ms, multi-band acceleration factor of 4, field of view of 206 mm, RL phase encoding direction, and 3.2 mm isotropic voxels). A T1 structural scan was first co-registered with a standard MNI template, followed by the co-registration of functional images to this aligned structural scan. The acquired images were analysed using fMRIprep. The pre-processing of the images consisted of slice-timing correction, realignment, spatial normalisation to the standard EPI template of the Montreal Neurological Institute (MNI), and spatial smoothing using a Gaussian kernel of 6-mm full-width at half maximum. Head motion was investigated for any excessive movement. 7 subjects were excluded where the head motion was large (mean frame-wise displacement greater than 0.5 (Parkes, 2018). Further information can be found in Supplementary Material

### Measurement of subjective effects

Subjective experiences were assessed using the retrospective Altered States of Consciousness (ASC) questionnaire completed 360 minutes following psilocybin administration. Behavioural changes under psilocybin were measured using subdimension scales that assessed the subjective experience of disembodiment (Studerus et al., 2010). The three questions used to measure disembodiment on this scale were: “I felt as if I no longer had a body”, “I had the impression I was out of my body” and “I felt as if I was floating”. See Supplementary Material for further details.

### Group selection and analysis conditions

Participants were selected for effective connectivity analysis based on their self-reported subjective effects during MRI scans. This selection included 44 subjects, divided based on their disembodiment scores from the ASC: disembodiment scores above 60, indicating strong disembodiment experience totalled 22 participants, mean = 83.68/100; range = 63-100, SD = 11.80, and 22 subjects with scores below 30, indicating mild disembodiment experience, mean = 8.41/100; range = 0-24, SD = 8.71. Both groups were analysed to assess the effects of psilocybin on brain connectivity during rest and music-listening conditions. We report the results for the high disembodiment group in the main text while the results for low disembodiment group are reported in the supplementary material.

### Dynamic causal modeling (DCM)

DCM estimates dynamic changes in brain activity, assessing how selected brain regions influence others through the generative modelling of the involved brain circuitry. DCM determines, using model selection procedures, the set of parameters best suited for a causal interpretation of timeseries data. We employed DCM in this study to elucidate the mechanistic, causal relationships among the selected brain regions, underlying the effects of psilocybin. DCM is capable of deciphering hierarchical connectivity patterns, which can only be inferred by modelling the directed connectivity, and provide a comprehensive framework for understanding the change and influence of brain network states. Specifically, we used spectral DCM, which is designed for resting state fMRI analysis (K. J. Friston, Kahan, Biswal, & Razi, 2014; Razi et al., 2015). Spectral DCM infers the effective connectivity that best explains the observed cross-spectral density, a data feature related to functional connectivity (Novelli, Friston, & Razi, 2023).

### Selection of model regions of interest (ROIs) and extraction of region coordinates across subjects

We investigated the effective connectivity of regions reported to be involved in the bodily sense of self and out of body experiences (Alexander et al., 2021; Borders, 2020; Underwood, Tolmeijer, Wibroe, Peters, & Mason, 2021). Our region of interest (ROI) coordinates were selected from previous clinical and experimental OBE research and were the bilateral AG (Blondiaux, Heydrich, & Blanke, 2021), TPJ (composed of the supramarginal gyrus (SMG), the superior temporal gyrus (STG) and sulcus (STS)), and AI (Ionta et al., 2011; Ionta et al., 2014). These literature-derived coordinates corresponded to regions in the AAL atlas and collectively spanned the TPJ while maintaining sufficient spatial separation to meet the requirements of dynamic causal modelling. We used 8mm spheres centred on these coordinates and were intersected with corresponding Automated Anatomical Labeling (AAL) regions (Tzourio-Mazoyer et al., 2002). The time series for each ROI was computed as the first principal component of the voxel activity within an 8 mm radius sphere centred on the ROI coordinates (as listed in Table 1).

**Table 1.** Coordinates of regions of interest.

| <u>Region</u> | <u>MNI coordinates</u> |  |  |
| --- | --- | --- | --- |
|  | *x* | *y* | *z* |
| Left Angular Gyrus | -63 | -56 | 26 |
| Right Angular Gyrus | 60 | -48 | 33 |
| Left Anterior Insula | -46 | 2 | 0 |
| Right Anterior Insula | 46 | 2 | 0 |
| Left Temporoparietal Junction | -54 | -30 | 19 |
| Right Temporoparietal Junction | 57 | -29 | 21 |

### Specification and inversion of DCM

A fully connected DCM model was specified for the six ROIs defined in Table 1, without any exogenous inputs. The DCM for each subject was then inverted using spectral DCM (K. J. Friston et al., 2014) to infer the effective connectivity that best explains the observed cross-spectral density for each subject. This procedure was repeated for each condition (resting state and music) and group (low and high OBE scores). The explained variance averaged 86.51% for the psilocybin rest condition and 84.92% for the psilocybin music condition, indicating a successful fit of the DCM model to the data.

### Second-level analysis using parametric empirical Bayes

The effective connectivity inferred by spectral DCM for each subject was taken to the second (group) level to test hypotheses concerning between-group effects. A Bayesian general linear model (GLM) was employed to decompose individual differences in effective connectivity into hypothesised group-average connection strengths and unexplained noise. Hypotheses on the group-level parameters are tested within the parametric empirical Bayes (PEB) framework (K. Friston, Zeidman, & Litvak, 2015; K. J. Friston et al., 2016), where both the expected values and the covariance of the parameters were considered. That is, precise parameter estimates influence the group-level result more strongly than uncertain estimates, which are down-weighted. Bayesian model reduction (BMR) is used as an efficient form of Bayesian model selection (K. Friston et al., 2015; K. J. Friston et al., 2016) to remove any connections that didn’t contribute to increase the model evidence. Please see the supplementary material for further technical details about PEB.

### Interpretation of Results

In DCM, effective connectivity is measured in the unit of Hertz (Hz). The unit of Hz quantifies the (inverse) rate of change in (neural states) per unit time that one brain region causes in another. Self-connections are log-scaled, to make the model estimation numerically stable, hence have no unit.

Behavioural associations express the strength of association between neural connectivity estimates and behavioural scores. These are the normalised beta (β) coefficients estimated when fitting the group level Bayesian GLM (i.e., the PEB model). A statistical threshold of posterior probability > .99 (equivalent to a very strong evidence) was applied to show the significance of relationship between observed neural patterns and measured behavioural outcomes.

Results reported here are for the high OBE group. Low OBE group results are found in the supplementary material.

## Results

The results show effective connectivity changes from baseline (no psilocybin) to the corresponding rest and music-listening scans under psilocybin, in participants reporting high and low disembodiment scores (Fig. 1). Disembodiment scores—used to group participants for comparative analysis—were assessed as alterations in bodily self-consciousness, measured at the end of the psilocybin session using the Altered States of Consciousness scale (see Methods). Including both scan types allowed us to identify connection changes that remained consistent across scans in participants with high disembodiment scores, as well as those that varied depending on context. Connections that changed consistently across rest and music scans in the high disembodiment group, and did not appear in either scan of the low disembodiment group, were retained to isolate effective connectivity changes that differentiate participants with high disembodiment. These are presented in Fig. 2 and Table 2. Mean effective connectivity at baseline (no psilocybin) is summarised in Supplementary Table S5.

**Fig 1.**
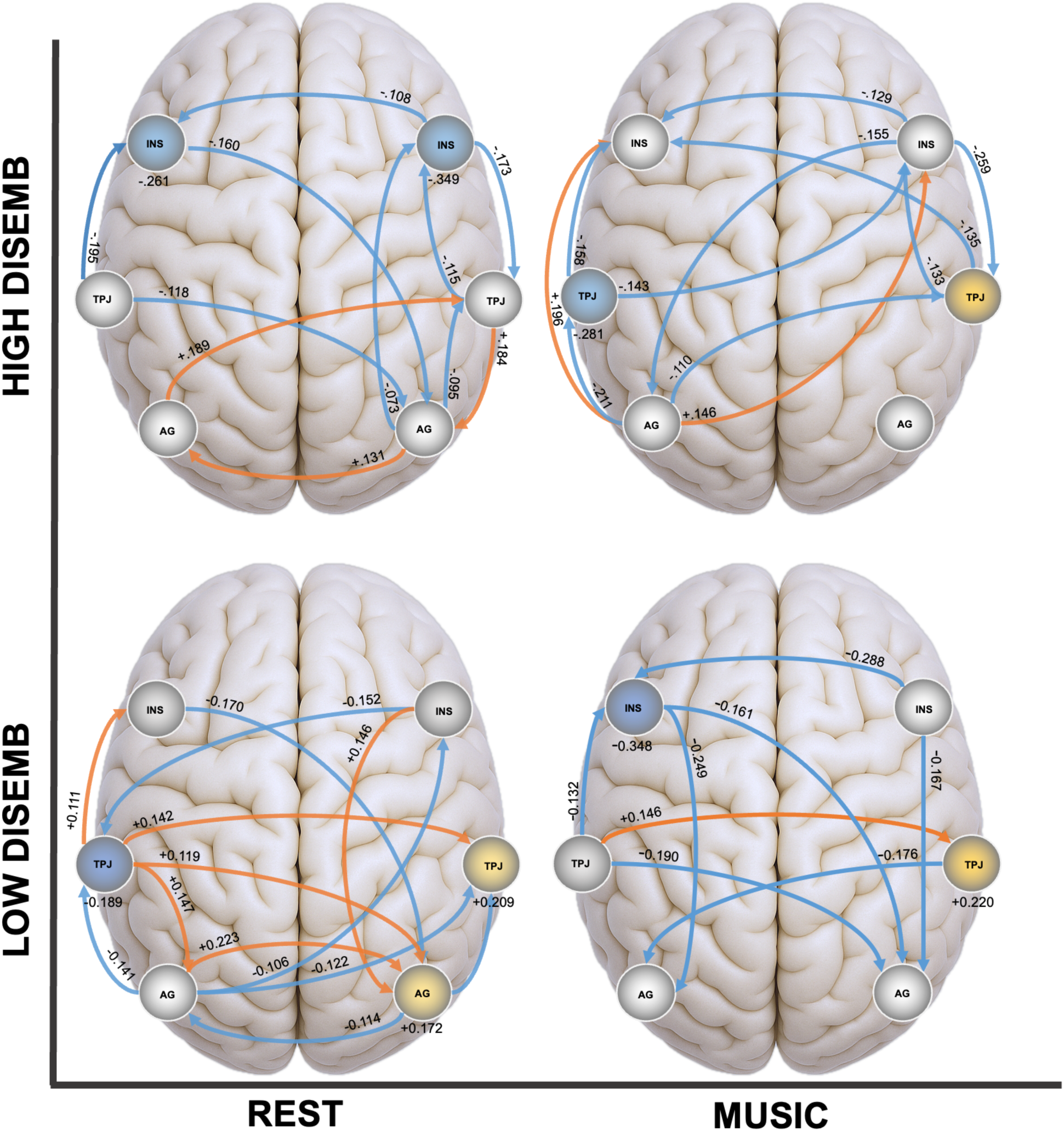
Effective connectivity changes from baseline to psilocybin 80 minutes post-administration for individuals with high and low disembodiment scores. Top left column is rest condition; right column is music condition; top row is high disembodiment; bottom row is low disembodiment. Connections show changes in effective connectivity compared to baseline (control). The shown values display effect sizes (called posterior expectations) of connections in Hz (except the inhibitory self-connections, which are log-scaled). Those connections and their associations not reported did not exceed the statistical threshold. The high disembodiment group is operationalised by score of disembodiment greater than 60 (out of 100). All results are for posterior probability > 0.99 (amounting to very strong evidence). Table of results for all conditions, including baseline mean effective connectivity, are found in Supplementary.

**Fig 2.**
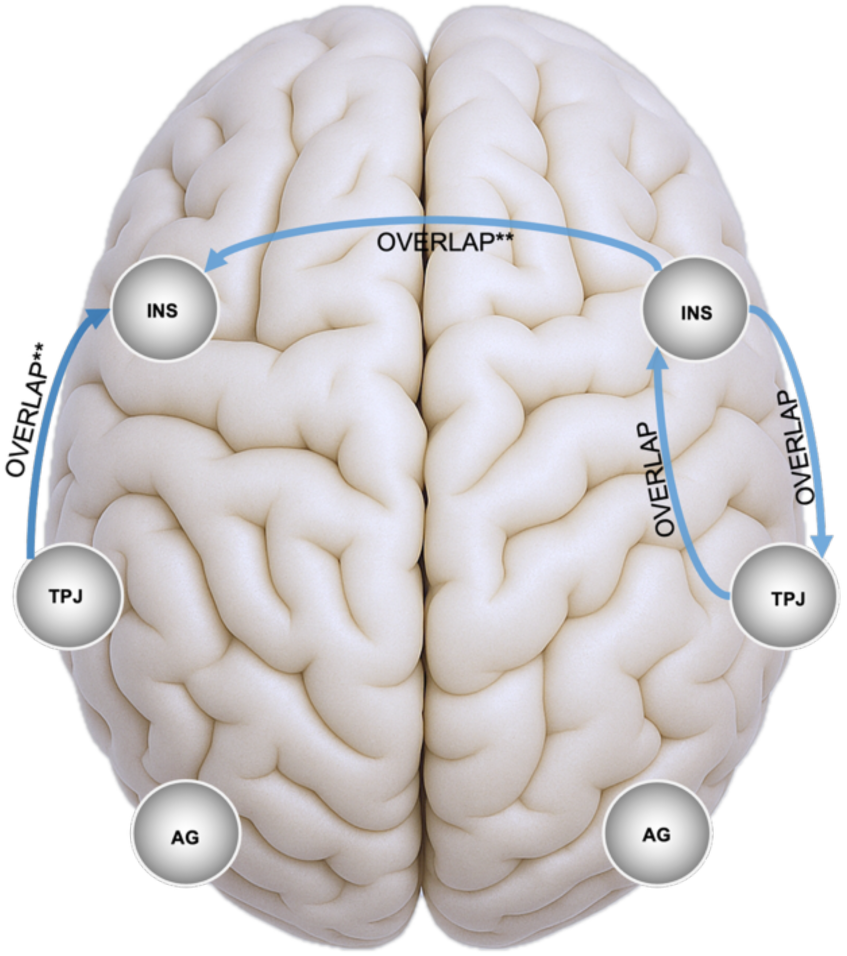
Effective connectivity changes for individuals with high disembodiment scores demonstrating overlapping connections (same valence) for resting state and music conditions. Double asterisks indicate the connection with same connection and valence was found in the low disembodiment group’s rest and or music scan. Change of valence in a connection between resting state and music scans are also demonstrated. All results are for connections exceeding posterior probability > 0.99 (amounting to very strong evidence).

**Table 2.** Changes in effective connectivity common to both rest and music scans under psilocybin in the high disembodiment group. These connections did not appear in either condition of the low disembodiment group. Normalised β coefficients are shown as effect sizes for posterior probability > 0.99. See Supplementary for full connection data and Table S5 for baseline values.

| *From* | *To* | *Rest Scan*<br>*Effect Size (Hz)* | *Music Scan*<br>*Effect Size (Hz)* |
| --- | --- | --- | --- |
| Right Anterior Insula | Right Temporoparietal Junction | -0.173 | -0.259 |
| Right Temporoparietal Junction | Right Anterior Insula | -0.115 | -0.133 |

### Effective connectivity changes under psilocybin in the high disembodiment group

In the rest scan, connectivity from and right AG to left AG demonstrated increased excitatory connectivity (0.131Hz) from baseline (no psilocybin). Similarly, the left AG to the right TPJ and right TPJ to right AG showed increased excitatory change during rest (0.189 and 0.184 Hz, respectively), changing these connections baseline mean effective connectivity from inhibitory (−0.181 and −0.124 Hz, respectively), to slightly excitatory (0.008 and 0.06 Hz, respectively). During the music scan, excitation in connections like those from the left AG to both the left and right AI was slightly elevated (0.196 and 0.146 Hz, respectively). Both rest and music scans showed consistent right TPJ to right AI inhibited connectivity (rest condition −0.115 Hz; music condition −0.133 Hz). We also found the opposite connection, right AI to right TPJ, showed a shift toward inhibition across both rest (−0.173 Hz) and music (−0.259 Hz). Connectivity patterns in the low disembodiment group are included in Fig. 1 to support contrastive interpretation. Full table of values for both groups and conditions can be found in Supplementary.

### Effective connectivity changes under psilocybin connections that overlap between rest and music scans for high disembodiment group

## Discussion

The results of our investigation into the effects of psilocybin on the TPJ demonstrate a notable pattern of inhibitory effective connectivity change (Fig 1). Among these findings, right TPJ to right AI inhibited connectivity was consistent across rest and music scans (rest condition (−0.115 Hz); music condition (−0.133 Hz). We also found the opposite connection, right AI to right TPJ, demonstrated a change towards inhibition across both rest (−0.173 Hz) and music (−0.259 Hz). Notably, the inhibition was greater during music (−0.259 Hz) than rest (−0.173 Hz), indicating that the disruption from right AI to right TPJ connectivity may be amplified by music stimulation under psilocybin in the high disembodiment group. This pattern of effective connectivity reduction, particularly involving the TPJ, underscores a robust inhibitory effect modulated by psilocybin and speaks to the function of TPJ and AI. The TPJ and AI are crucial for integrating sensory and spatial information necessary for body representation. Finding these connections associated with experienced intensity of disembodiment under psilocybin across both scans and exclusive to the high disembodiment group suggests their inhibitory change as neural mechanisms of OBEs (see Fig 2 and Table 2).

### Music modulation of effective connectivity change

We identified a subset of connections exhibiting what may be considered *synergistic effects*, meaning they changed from rest to music only in the context of high disembodiment—for example, increased excitatory connectivity from the left AG to the left and right AI. Connections estimated across both scans in the high disembodiment group that were either right-lateralised or originated from the right hemisphere showed greater inhibitory change from rest to music. For instance, the connection from the right AI to the right TPJ shifted from −0.173 Hz at rest to −0.259 Hz during music, while its reciprocal connection—from the right TPJ to the right AI—shifted from −0.115 Hz at rest to −0.133 Hz during music. This enhanced inhibition may reflect the synergistic effects of music stimulation under psilocybin (Barrett, Preller, & Kaelen, 2018), specific to participants reporting high disembodiment scores.

Although the music scan took place 7 minutes after the rest scan, natural variability in psilocybin onset and peak timing across participants makes delayed peak effects an unlikely explanation for the observed differences between conditions. Importantly, inhibitory changes in the right AI to left AI and left TPJ to left AI connections were also observed during the music condition in both low and high disembodiment groups. This suggests that some aspects of TPJ–insula connectivity may reflect a general interaction between music and psilocybin, independent of subjective disembodiment.

### AG connections

TPJ connectivity with other regions also showed changes in the high disembodiment group, which were specific to the scanning sequence. Interestingly connections from AG accounted for four out of five excitatory changes found in our results for the high disembodiment group. Functional imaging has previously found AG involvement in processing spatial orientations and perspectives that are distorted during OBE experiences (De Ridder, Van Laere, Dupont, Menovsky, & Van de Heyning, 2007). Electrical stimulation targeting the right angular gyrus induced perceptual distortions including out-of-body experiences, confirming the role of the angular gyrus within the broader context of the temporo-parietal junction in such phenomena (Blanke, Ortigue, Landis, & Seeck, 2002). This direct targeting provided evidence of the angular gyrus’s involvement in integrating sensory and vestibular information critical for maintaining normal self-body perception. Our findings support the associations between alteration of this connectivity and experiences of disembodiment, suggesting modulation of excitation of specific AG connectivity by context as a factor in OBEs.

For reference and comparison, we analysed a group of participants with low disembodiment scores, operationalised as disembodiment scores less than 30 (out of 100). The low disembodiment group demonstrated an array of connectivity changes in our model of the TPJ, with fewer patterns of inhibition compared to those in the high disembodiment group, particularly in the resting state condition. The reduced pattern of inhibition change may reflect less disrupted sensory integration processes, potentially correlating with fewer or less intense disembodiment experiences. The changes to effective connectivity in this group showed no pattern of inhibition between the right TPJ and right AI.

Observing greater inhibition during the resting-state condition in participants reporting high disembodiment, alongside more excitatory changes in those reporting low disembodiment, suggests that a generalised downregulation of TPJ connectivity—particularly involving specific AI-TPJ inhibitory changes localised to the right hemisphere—may play a central role in the intensity of disembodiment. This alteration in connectivity aligns with previous clinical and experimental findings (Blanke et al., 2002; Blondiaux et al., 2021; De Ridder et al., 2007; Ionta et al., 2011; Ionta et al., 2014) and adds compelling evidence for a shared neural basis of disembodiment across both spontaneous and induced states.

While Blondiaux et al. mapped trait-level disembodiment using lesion and functional connectivity data, and Blanke (2002) and De Ridder et al. (2007) reported acute state-level disembodiment during direct cortical stimulation, Ionta et al. demonstrated moment-to-moment fluctuations in disembodiment using robot-supported virtual reality manipulations in healthy participants. In contrast, our study captures a sustained, pharmacologically-induced disembodiment state, offering a unique window into the network dynamics underlying prolonged self-detachment.

By identifying specific inhibitory interactions between the TPJ and insula under psilocybin, our findings strengthen the emerging view that this circuit plays a central role in maintaining coherent bodily self-location. These results support a model in which psilocybin disrupts the integration of sensory information through decreased connectivity within key sensory integration hubs—including the TPJ and insula—and highlight the generalisability of these mechanisms across psychedelic, clinical, and experimental contexts.

Psilocybin primarily exerts its effects through its active metabolite, psilocin, which agonises serotonin receptors, notably the 5-HT2A receptor (Nichols, 2016). This receptor, prevalent in cortical regions, is pivotal in modulating neurotransmission and connectivity (Hensler, 2012). Activation of the 5-HT2A receptor by psilocin is believed to enhance neural plasticity, potentially facilitating a temporary reorganisation of functional neuronal connectivity (Calder & Hasler, 2023; Lord et al., 2018). Understanding the reorganisation is vital to deduce the connectivity responsible for features of consciousness alteration. Psilocin’s interaction with serotonin receptors are a likely mechanism underlying the shift in excitatory-inhibitory balance within neural circuits, observed here and in previous work as the reductions and increases in effective connectivity (Stoliker, Novelli, et al., 2022; Devon Stoliker et al., 2024; D. Stoliker et al., 2024). For instance, psilocybin could augment glutamatergic transmission in various brain regions, while simultaneously boosting local inhibitory feedback (Mason et al., 2020), resulting in complex, region-specific alterations in directed connectivity.

The role of serotonin in brain function includes mood, perception and cognitive functions such as memory and attention, mediated by various brain regions, including the temporal and parietal lobes. Previous research indicates that psychedelics impact sensory processing and gating—mechanisms that modulate and filter sensory information (Preller et al., 2019; Swanson, 2018). By disrupting the usual sensory gating via serotonin receptors, particularly affecting TPJ sensory integration, psilocybin may blur the perceptual boundaries between the self and the environment, contributing to experiences like disembodiment. Clinical and experimental evidence of TPJ function in OBEs emphasise its significance and suggests extensive possibilities to research psychedelic effects by investigating connectivity localised and processed through the TPJ. Furthermore, the interaction of external stimuli, such as music, with serotonin-mediated pathways also modulated observed connectivity patterns, consistent with previous findings (Mediano et al., 2024) (Stoliker et al., 2025). Our findings on the differences between rest and music conditions between high and low disembodiment groups provide indications of specific stimuli-modulated connections under psilocybin.

Understanding how psilocybin disrupts normal sensory processing leading to altered perceptions is explored by several works through the theoretical framework of predictive coding, a theory that captures the nuanced interplay between top-down expectations and bottom-up sensory inputs (K. Friston & Kiebel, 2009) (Stoliker, Egan, Friston, et al., 2022) (Pink-Hashkes, Rooij, & Kwisthout, 2017) (Carhart-Harris & Friston, 2019). This theory outlines a hierarchical organisation of top-down predictions and bottom-up prediction errors and may be particularly applicable to the distinction of signals in regions like the TPJ and insula, which integrate multiple sensory inputs to forge a coherent sense of self. From the view of predictive coding, changes to our model of the TPJ suggest a complex interaction of multisensory organisation.

Changes in the precision of sensory and high-level predictions are suggested to underlie experiences of ego dissolution, where individuals report a blurring of the boundary between self and environment (Stoliker, Egan, & Razi, 2022). This phenomenon, reported under the influence of substances like LSD or psilocybin, is relevant to the integration of the temporoparietal junction (TPJ) with cognitive brain regions. The TPJ is a crucial component of the Default Mode Network (DMN), which is commonly associated with self-related processing. The dissociation with the sense of self and reduced separation from the environment during psychedelic ego dissolution experiences suggests a possible influence of change to the organisation and integration of interoceptive and exteroceptive signals. However, the abundance of interest in ego dissolution has largely overlooked the central importance of the TPJ to its phenomenology, suggesting the value in understanding the role of TPJ in OBEs and its associations to the therapeutic benefits of psychedelics. For example, the transformative effects reported from spontaneous OBEs and attitudes towards dying, aligns with the therapeutic benefits observed in ego dissolution states, wherein disembodiment may have therapeutic importance. Specifically, the similarities between near-death experiences (NDEs), which can result in transformative therapeutic changes (Sweeney et al., 2022), OBE-induced transformations (Shaw, Gandy, & Stumbrys, 2023), and psychedelic-induced transformations (Martial et al., 2021) indicate common phenomenology that merit exploring similarities in brain connectivity changes. This line of research can also be extended to understanding the role of OBEs in trance states (Hove et al., 2016), suggesting broad implications for OBE research and potential therapeutic applications.

### Limitations

The difficulty in describing the function of any given circuit is widespread in neuroscience, stemming from the multiple roles ascribed to each brain circuitry. This complexity poses significant challenges in developing models of effective connectivity, which must simplify the brain’s highly integrated nature for modelling and interpretation based on prior hypotheses. Although these models offer valuable insights, they are approximations and should be viewed as complementary to broader research. Our study also faced variability in psychedelic experiences among participants, particularly in the timing and intensity of disembodiment measures, which relied on a 19mg dose of psilocybin. Not all subjects in the high disembodiment group experienced full disembodiment (See Methods), limiting the generalisability of our findings and highlighting the complexity of capturing the nuanced effects of psychedelics, which extend beyond localised brain regions such as the TPJ. Another limitation is that psychedelic subjective effects covary. The co-occurrence of diverse psychedelic effects and disembodiment complicates the specificity of our findings, as psychedelic-induced disembodiment cannot be definitively isolated from broader experiential effects. While our grouping approach contrasted participants based on disembodiment intensity, the connectivity changes observed may still reflect overlapping dimensions of the psychedelic state. However, given the robust prior literature implicating the TPJ in OBEs, we interpret these findings as consistent with known functional roles of this region in bodily self-location and disembodiment. Lastly, the reliance on self-reported Altered States of Consciousness (ASC) Likert scales may introduce subjectivity and potential recall bias.

### Future directions

A promising approach to extend the generalisability of our findings is to investigate case studies of profound OBEs. Future experiments may also be designed to compare the effects of eyes-open vs eyes-closed conditions in modulating disembodiment experiences and underlying connectivity, following evidence of a reducing effect of eyes-open on psychedelic effects that may limit OBEs has been reported (Mediano et al., 2024; Stoliker et al., 2025). Such comparative approaches could help in differentiating the neurophysiological signatures of disembodiment specific to psychedelic use and advance our understanding of the mechanisms by which psychedelics alter consciousness and perception.

Further investigations could explore the influence of contextual elements, such as mindset and setting, on OBE experiences. Additionally, combining psilocybin administration with established OBE induction techniques—such as those utilising virtual reality (VR) to manipulate bodily self-consciousness—may provide deeper insights into the neural mechanisms underlying disembodiment. For instance, Lenggenhager et al. (2007) (Lenggenhager, Tadi, Metzinger, & Blanke, 2007) demonstrated that synchronous visuo-tactile stimulation in a VR setup can induce OBE-like sensations in healthy individuals. Implementing such paradigms before and after psilocybin administration could help delineate the specific contributions of pharmacological and contextual factors to disembodiment experiences. Moreover, assessing disembodiment scores in relation to these combined interventions may clarify the temporal dynamics and subjective intensity of OBEs under the influence of psychedelics.

Our study’s inclusion of both a resting-state and a music condition marks a step toward understanding the effects of context. However, more comprehensive studies involving varied rituals—such as shamanic ceremonies that combine psychedelic ingestion with fasting, prolonged dancing, and extended singing and chanting—could offer deeper insights into how contextual factors modulate brain responses. Investigating clinical or experimental datasets of OBEs using DCM may also provide valuable points of comparison, helping to distinguish psychedelic-induced OBEs from those that occur spontaneously or in clinical settings. Finally, integrating technologies such as high-definition transcranial direct current stimulation (HD-tDCS) with psychedelic use may further prime key neural regions implicated in OBEs. These approaches could enhance our understanding of connectivity changes in the TPJ and related areas during induced OBEs, advancing efforts to uncover the neural underpinnings of these profound alterations in self-consciousness.

### Conclusion

Psychedelics, through 5-HT receptor agonism, alter bodily ownership and self-location in ways that parallel neurological disruptions and experimentally induced illusions. Psilocybin provides a controlled method for inducing disembodiment, allowing tests of its similarity to clinical and spontaneous OBEs at the level of neuronal connectivity. Using effective connectivity analyses, we demonstrated that psilocybin induces inhibitory changes to right TPJ connectivity, aligning with disruptions reported in non-pharmacological contexts. These findings support a shared mechanism underlying OBEs across altered states. Future work should disentangle the diverse subjective effects of psychedelics and their influence on regional and network-level connectivity involving the TPJ, mapping how these changes alter sense of bodily self and subject–object perspective. Advancing this understanding holds value for both clinical interventions and the science of consciousness.

## Data Availability

All data produced in the present study are available upon reasonable request to the authors

## Acknowledgments and Disclosures

All data are available in the main text, Supplementary Material, or by request to the corresponding author. The authors report no biomedical financial interests or potential conflicts of interest.

This article, or part of it, has been posted on medRxiv doi:

## Author contributions

Conceptualisation: DS, AR, OB, FB

Methodology: DS, AR

Investigation: DS, AR

Visualisation: DS

Writing—original draft: DS

Writing—review & editing: DS, AR, OB, FB

