## Supplementary Material for "Psilocybin Modulates TPJ Effective Connectivity during Out-of-Body Experiences"

Devon Stoliker, Monash Biomedical Imaging, 762-772 Blackburn Rd, Clayton VIC 3168. Australia.

+ Joint Senior Authors

### 1.1 Spectral Dynamic Causal Modelling

Dynamic causal modelling (DCM) is Bayesian framework that infers the directed (causal) connectivity among the neuronal systems – referred to as effective connectivity. We recently proposed a new DCM for resting state fMRI – based upon a deterministic model that generates predicted cross spectra – referred to as spectral DCM. In order to model resting state activity – in the absence of external stimuli – we will have to add a stochastic component, i.e. neural fluctuations, to the classical DCM based on ordinary differential equations. Mathematically, we can express the formulation of the stochastic generative model using a set of two equations. First is the neuronal state equation, namely

$$\dot{x}(t) = f(x(t), u(t), \theta) + v(t), \qquad \text{(S1)}$$

and second is the observation equation, which is a static nonlinear mapping from the hidden physiological states in (1) to the observed BOLD activity and is written as:

$$y(t) = h(x(t), \varphi) + e(t), \tag{S2}$$

where $\dot{x}(\mathrm{t})$ is the rate of change of the neuronal states $x(t)$, $\theta$ are unknown parameters (i.e. the effective connectivity) and $v(t)$ (resp. $e(t)$) is the stochastic process – called the state noise (resp. the measurement or observation noise) – modelling the random neuronal fluctuations that drive the resting state activity. In the observation equations, $\varphi$ are the unknown parameters of the (haemodynamic) observation function and $u(t)$ represents any exogenous (or experimental) inputs that drive the hidden states – that are usually absent in resting state designs (1). Spectral DCM furnishes a constrained inversion of the stochastic model by parameterising the neuronal fluctuations $v(t)$. Spectral DCM simplifies the generative model by replacing the original timeseries with their second-order statistics (i.e., cross spectra). This means, instead of estimating time varying hidden states, we are estimating their covariance which is time invariant. Then we simply need to estimate the covariance of the random fluctuations; where a scale free (power law) form for the state noise (resp. observation noise) is used – motivated from previous work on neuronal activity (2-4) – as follows:

$$g_v(\omega, \theta) = \alpha_v \omega^{-\beta_v}$$

$$g_e(\omega, \theta) = \alpha_e \omega^{-\beta_e} \tag{S3}$$

Here, $\{\alpha, \beta\} \subset \theta$ are the parameters controlling the amplitudes and exponents of the spectral density of the neural fluctuations. The parameterisation of endogenous fluctuations means that the states are no longer probabilistic; hence the inversion scheme is significantly simpler, requiring estimation of only the parameters (and hyperparameters) of the model.

We used standard Bayesian model inversion to infer the parameters of the model in (1), (2) and (3), from the observed signal $y(t)$. The description of the Bayesian model inversion procedures based on variational Laplace can be found elsewhere for the interested readers (5-7).

### 1.2 Parametric Empirical Bayes

Empirical Bayes refers to the Bayesian inversion or fitting of hierarchical models. In hierarchical models, constraints on the posterior density over model parameters at any given level are provided by the level above. These constraints are called empirical priors because they are informed by empirical data. We recently introduced a second-level or between-subjects model over parameters, which represents how individual (within-subject) connections derive from the subjects' group membership (8) – based on parametric empirical Bayes (PEB). This approach calls on Bayesian Model Reduction (BMR) to finesse the inversion of multiple models of a single dataset or a single (hierarchical) model of multiple datasets. BMR allows one to compute posterior densities over model parameters, under new prior densities, without explicitly inverting the model again. For example, one can invert a DCM for each subject in a group and then evaluate the posterior density over group effects, using the posterior densities over parameters from the single subject inversion. This may improve subject-specific parameter estimates, by using group-level estimates to rescue individual DCM from local optima. Mathematically, for DCM studies with *N* subjects and *M* parameters per DCM, we have a hierarchical model, where the responses of the *i*-th subject and the distribution of the parameters over subjects can be modeled as:

$$y_i = \Gamma_i^{(1)}(\theta^{(1)}) + \varepsilon_i^{(1)} \qquad \text{(S4)}$$

$$\theta^{(1)} = \Gamma^{(2)}(\theta^{(2)}) + \varepsilon^{(2)}$$

$$\theta^{(2)} = \eta + \varepsilon^{(3)}$$

where, $y_i$ is the BOLD time series from *i-th* subject *and* $\Gamma_i^{(1)}$ is a nonlinear mapping from the parameters of a model to the predicted response $y$ for e.g. as shown in Eq. S1 above. $\varepsilon_i^{(1)}$ is independent and identically distributed (i.i.d.) observation noise (equivalent to $e(t)$ in Eq. S2). In this hierarchical form, *empirical priors* encoding second (between-subject) level effects place constraints on subject-specific parameters. The second level would be a linear model where the random effects are parameterised in terms of their precision:

$$\Gamma^{(2)}(\theta^{(2)}) = (X \otimes W)\beta$$

where, $\beta \subset \theta$ are group means or effects encoded by a design matrix with between $X$ and within-subject $W$ parts. The between-subject part encodes differences among subjects or covariates such as age, while the within-subject part specifies mixtures of parameters that show random effects. We assume that the first column of the design matrix is a constant term, modelling group means and subsequent columns encode group differences or covariates such as age.

### 1.3 Self-connections in DCM

Please note that in DCM, the self-connections are always modelled as inhibitory (to preclude any run-away excitation), but these parameters in the model are log-scaled for the sake of numerical stability of the model fitting procedures. This (log) scaling means that these self-connections can take both positive (red) and negative values (blue). A positive self-connection means a relative increased inhibition, whereas a negative self-connection means a relative decreased inhibition (i.e., disinhibition). Inhibitory self-connections control the regions' gain or sensitivity to inputs. Only the self-connections are log-scaled in DCM.

### 1.4 Subjective Effects (5D-ASC)

Disembodiment was measured on the retrospective Altered States of Consciousness self-report questionnaire (ASC) 80 minutes after the oral administration of 19mg psilocybin and scored between 1-100 on a Likert scale.

The ASC disembodiment scale consisted of the following three questions: "I felt as if I no longer had a body", "I had the impression I was out of my body" and "I felt as if I was floating".

The 22 participants categorised into the high disembodiment group level averaged = 83.68/100; range = 63-100, SD = 11.80; the 22 participants categorised into the low disembodiment group level averaged = 8.41/100; range = 0-24, SD = 8.71

mean = 83.68/100; range = 63-100, SD = 11.80, and 22 subjects with scores below 30, indicating mild disembodiment experience, mean = 8.41/100; range = 0-24, SD = 8.71.

### 1.5 Participants

All participants were deemed healthy after screening for medical history, physical examination, blood analysis, and electrocardiography. Participants were asked to abstain from prescription and illicit drug use two weeks prior to first testing and throughout the duration of the study and abstain from alcohol use 24 hours prior to testing days. Urine tests and self-report questionnaires were used to verify the absence of drug and alcohol use. Urine tests were also used to exclude pregnancy. Further exclusion criteria included poor knowledge of the English language, cardiovascular disease, history of head injury or neurological disorder, history of alcohol or illicit drug dependence, MRI exclusion criteria, including claustrophobia, and previous use of a hallucinogenic drug. All participants provided written informed consent statements in accordance with Monash University Ethics Committee Guidelines. Subjects received written and oral descriptions of the study procedures, as well as details regarding the effects and possible risks of drug treatment.

### MRI Data Acquisition

Structural MRI data was acquired using a Siemens 3 Tesla Magnetom Skyra scanner at Monash Biomedical Imaging, Monash University, Australia. T1-weighted (T1w) anatomical images were obtained for each participant during two sessions: at baseline (no-psilocybin) and on the psilocybin administration day. The images were acquired using a 3D magnetisation-prepared rapid gradient-echo (MP-RAGE) sequence with a 32-channel head coil. The acquisition parameters were as follows: repetition time (TR) of 2300 ms, echo time (TE) of 2.07 ms, 192 slices per slab, 1 mm slice thickness, and 1 mm isotropic voxel size. The parallel acquisition technique mode used was GRAPPA, with an acceleration factor (PE) of 2 on the baseline day and 3 on the psilocybin administration day. A T2-weighted anatomical image was obtained only during the baseline session. The acquisition parameters were: TR of 3200 ms, TE of 452 ms, 176 slices per slab, 1 mm slice thickness, and 1 mm isotropic voxel size, with an acceleration factor of 2. Blood-oxygenation-level-dependent (BOLD) fMRI data was collected using a multi-echo, multi-band, echo-planar imaging, T2*-weighted sequence. The acquisition parameters were: TR of 910 ms, multi-echo TE of 12.60 ms, 29.23 ms, 45.86 ms, 62.49 ms, multi-band acceleration factor of 4, field of view of 206 mm, RL phase encoding direction, and 3.2 mm isotropic voxels. The scan durations were: resting state with eyes closed (8 minutes, 505 volumes), audio-guided meditation with eyes closed (6:30 minutes, 405 volumes), music listening with eyes closed (11:24 minutes, 728 volumes), movie watching (6:00 minutes, 372 volumes). The structural and functional MRI images acquired from the Siemens scanner were converted into the Neuroimaging Informatics Technology Initiative (NIfTI) format.

**Figure S1.**

$$\begin{bmatrix} 1 & 0 \\ 1 & 0 \\ 1 & 1 \\ 1 & 1 \end{bmatrix}$$

**Design matrix.** Design matrix and effective connectivity posterior expectation matrices are demonstrated. Designated the baseline group to serve as the baseline. Regressors in change design matrix encode: 1) baseline group 2) the additive effect of being in the second group (psilocybin after 80 min) relative to the placebo group.

### Table S1. Effective connectivity change under psilocybin for high disembodiment group

| ***Rest Scan***<br>***From*** | ***To*** | **Effect Size (Hz)** | **CI Low** | **CI High** |
|---|---|---|---|---|
| Left Angular Gyrus | Right Temporoparietal Junction | 0.189 | 0.092 | 0.285 |
| Right Angular Gyrus | Left Angular Gyrus | 0.131 | 0.052 | 0.21 |
| Right Angular Gyrus | Right Anterior Insula | -0.073 | -0.153 | 0.007 |
| Right Angular Gyrus | Right Temporoparietal Junction | -0.095 | -0.305 | -0.084 |
| Left Anterior Insula | Right Angular Gyrus | -0.160 | -0.266 | -0.053 |
| Left Anterior Insula | Left Anterior Insula | -0.261 | -0.433 | -0.089 |
| ***Right Anterior Insula*** | ***Left Anterior Insula*** | ***-0.108*** | -0.247 | 0.031 |
| Right Anterior Insula | Right Anterior Insula | -0.349 | -0.531 | -0.167 |
| ***Right Anterior Insula*** | ***Right Temporoparietal Junction*** | ***-0.173*** | -0.304 | -0.043 |
| Left Temporoparietal Junction | Right Angular Gyrus | -0.118 | 0.092 | 0.285 |
| ***Left Temporoparietal Junction*** | ***Left Anterior Insula*** | ***-0.195*** | -0.305 | -0.084 |
| Right Temporoparietal Junction | Right Angular Gyrus | 0.184 | 0.094 | 0.273 |
| ***Right Temporoparietal Junction*** | ***Right Anterior Insula*** | ***-0.115*** | -0.211 | -0.02 |
| ***Music Scan***<br>***From*** | ***To*** | **Effect Size (Hz)** | **CI Low** | **CI High** |
| **Left Angular Gyrus** | Left Anterior Insula | 0.196 | 0.096 | 0.296 |
| Left Angular Gyrus | Right Anterior Insula | 0.146 | 0.064 | 0.229 |
| Left Angular Gyrus | Left Temporoparietal Junction | -0.211 | -0.287 | -0.135 |
| Left Angular Gyrus | Right Temporoparietal Junction | -0.11 | -0.172 | -0.048 |
| Right Anterior Insula | Left Angular Gyrus | -0.155 | -0.24 | -0.07 |
| ***Right Anterior Insula*** | ***Left Anterior Insula*** | ***-0.129*** | -0.253 | -0.006 |
| ***Right Anterior Insula*** | ***Right Temporoparietal Junction*** | ***-0.259*** | -0.367 | -0.152 |
| ***Left Temporoparietal Junction*** | ***Left Anterior Insula*** | ***-0.158*** | -0.281 | -0.035 |
| Left Temporoparietal Junction | Right Anterior Insula | -0.143 | -0.233 | -0.053 |
| Left Temporoparietal Junction | Left Temporoparietal Junction | -0.281 | -0.423 | -0.139 |
| Right Temporoparietal Junction | Left Anterior Insula | -0.135 | -0.223 | -0.047 |
| ***Right Temporoparietal Junction*** | ***Right Anterior Insula*** | ***-0.133*** | -0.238 | -0.029 |
| Right Temporoparietal Junction | Right Temporoparietal Junction | 0.245 | 0.092 | 0.399 |

*Table S1. Extended table of Change of effective connectivity under psilocybin for high disembodiment group. Connections common to both rest and music scan are in bold italics. B coefficients (posterior probabilities) are listed as effect size for posterior probability .99. CI = confidence interval. See Supplementary further detail. Figures from main text below for reference.*

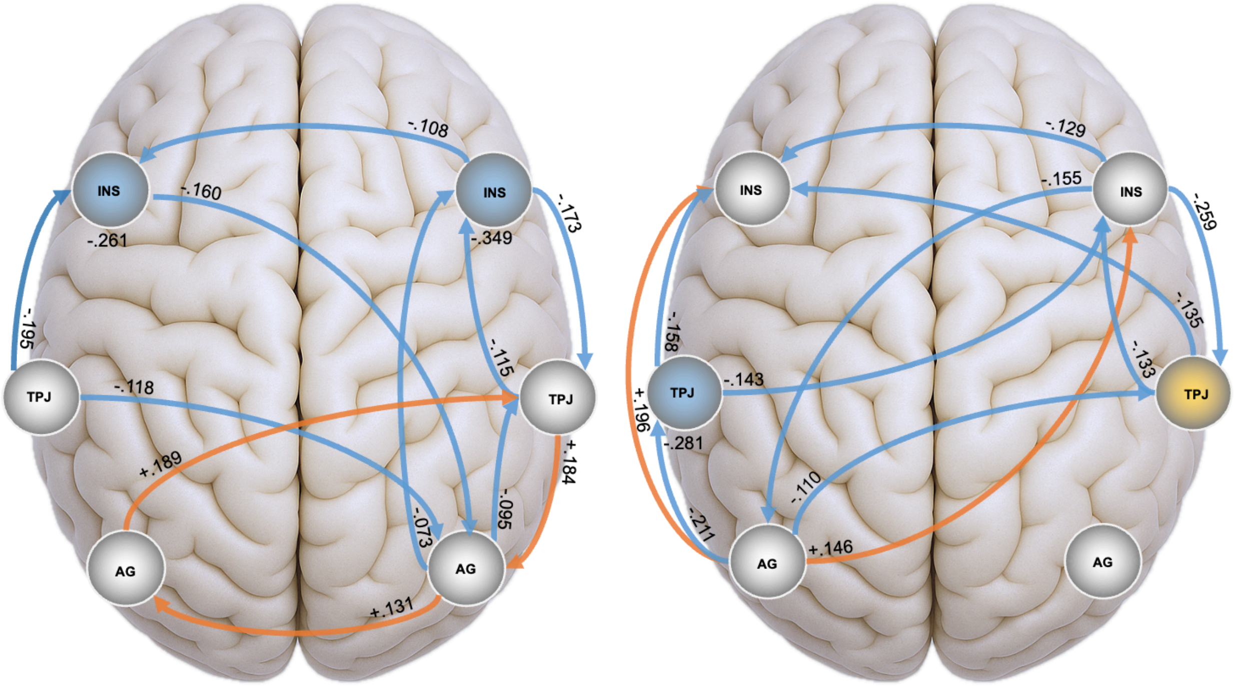

-.108
INS
-.160
INS
-.173
-.261
-.349
-.195
-.115
TPJ
-.118
TPJ
+.184
+.189
-.095
-.073
AG
AG
+.131
-.129
INS
-.155
INS
-.259
-.135
-.158
-.133
TPJ
-.143
TPJ
+.196
-.281
-.110
-.211
AG
+.146
AG

**Table S2.**

| ***Rest Scan*** | | | | |
|---|---|---|---|---|
| ***From*** | ***To*** | **Effect Size (Hz)** | **CI Low** | **CI High** |
| Left Angular Gyrus | Right Angular Gyrus | 0.223 | 0.155 | 0.292 |
| Left Angular Gyrus | Right Anterior Insula | -0.106 | -0.206 | -0.007 |
| Left Angular Gyrus | Left Temporoparietal Junction | -0.122 | -0.183 | -0.06 |
| Left Angular Gyrus | Right Temporoparietal Junction | -0.141 | -0.212 | -0.07 |
| Right Angular Gyrus | Left Angular Gyrus | -0.114 | -0.19 | -0.037 |
| Right Angular Gyrus | Right Angular Gyrus | 0.172 | 0 | 0.345 |
| ***Left Anterior Insula*** | ***Right Angular Gyrus*** | ***-0.170*** | -0.281 | -0.059 |
| Right Anterior Insula | Right Angular Gyrus | 0.146 | 0.013 | 0.279 |
| Right Anterior Insula | Left Temporoparietal Junction | -0.152 | -0.273 | -0.03 |
| Left Temporoparietal Junction | Left Angular Gyrus | 0.147 | 0.021 | 0.272 |
| Left Temporoparietal Junction | Right Angular Gyrus | 0.119 | 0.021 | 0.218 |
| Left Temporoparietal Junction | Left Anterior Insula | 0.111 | 0.017 | 0.205 |
| Left Temporoparietal Junction | Left Temporoparietal Junction | -0.189 | -0.372 | -0.007 |
| ***Left Temporoparietal Junction*** | ***Right Temporoparietal Junction*** | ***0.142*** | 0.034 | 0.249 |
| ***Right Temporoparietal Junction*** | ***Right Temporoparietal Junction*** | ***0.209*** | 0.044 | 0.375 |
| ***Music Scan*** | | | | |
| ***From*** | ***To*** | **Effect Size (Hz)** | | |
| Left Insula | Left Angular Gyrus | -0.249 | -0.349 | -0.149 |
| ***Left Insula*** | ***Right Angular Gyrus*** | ***-0.161*** | -0.289 | -0.034 |
| Left Insula | Left Insula | -0.348 | -0.5 | -0.195 |
| Right Insula | Right Angular Gyrus | -0.167 | -0.295 | -0.039 |
| Right Insula | Left Insula | -0.288 | -0.435 | -0.14 |
| Left Temporoparietal Junction | Right Angular Gyrus | -0.190 | -0.313 | -0.068 |
| Left Temporoparietal Junction | Left Insula | -0.132 | -0.265 | 0 |
| ***Left Temporoparietal Junction*** | ***Right Temporoparietal Junction*** | ***0.146*** | 0.048 | 0.244 |
| Right Temporoparietal Junction | Left Angular Gyrus | -0.176 | -0.252 | -0.099 |
| ***Right Temporoparietal Junction*** | ***Right Temporoparietal Junction*** | ***0.220*** | 0.077 | 0.363 |

*Table S2. Change of effective connectivity under psilocybin for low disembodiment group. Connections common to both rest and music scan are in bold italics. B coefficients (posterior probabilities) are listed as effect size for posterior probability .99. CI = confidence interval. See Supplementary further detail.*

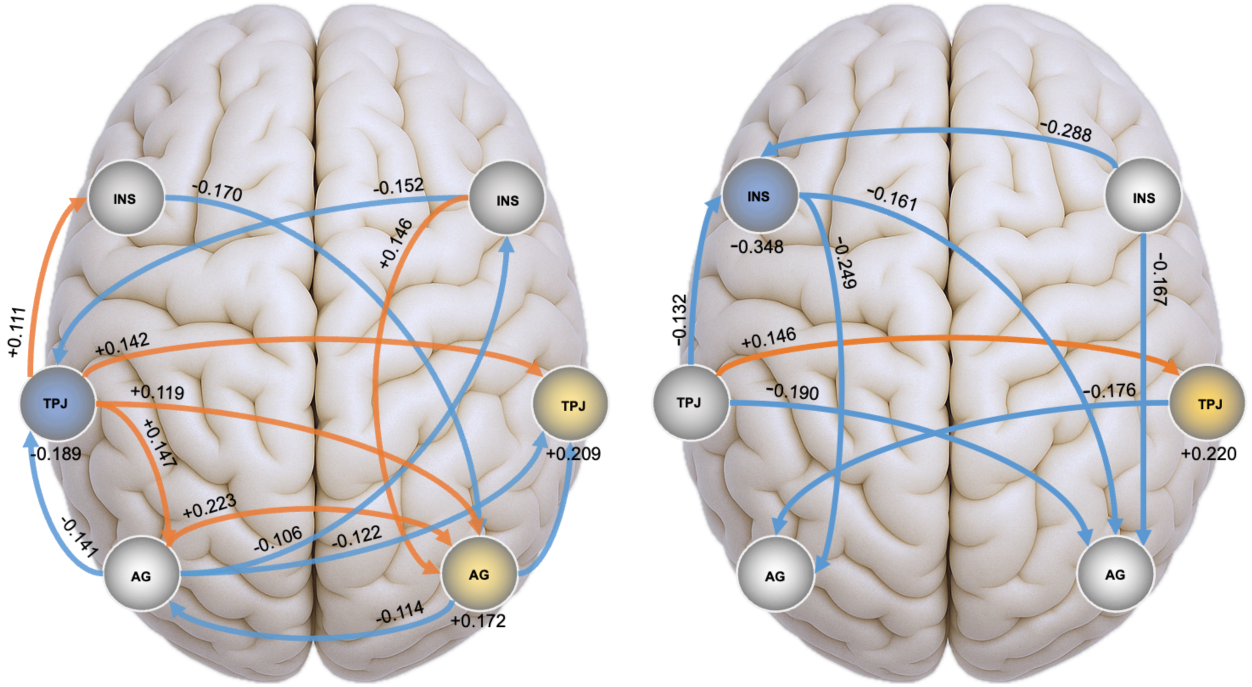


### Table S3.

| ***Low Disembodiment*** ***From*** | ***To*** | ***Rest Scan*** ***Effect Size (Hz)*** | ***Music Scan*** ***Effect Size (Hz)*** |
|---|---|---|---|
| Left Anterior Insula | Right Angular Gyrus | -0.170 | -0.161 |
| Left Temporoparietal Junction | Right Temporoparietal Junction | 0.142 | 0.146 |
| Right Temporoparietal Junction | Right Temporoparietal Junction | 0.209 | 0.220 |

*Table S3. Change of effective connectivity common to both rest and music scan under psilocybin for high disembodiment group. B coefficients (posterior probabilities) are listed as effect size for posterior probability .99. See Supplementary further detail.*

### Table S4.

| **From** | **To** | **High** Music | **High** Rest | **Low** Music | **Low** Rest |
|---|---|---|---|---|---|

| | | | | | |
|---|---|---|---|---|---|
| Left Temporoparietal Junction | Left Anterior Insula | -0.158 | -0.195 | -0.132 | 0.111 |

*Table S4. Change of effective connectivity valence change between high and low disembodiment group. B coefficients (posterior probabilities) are listed as effect size for posterior probability .99.*

### Table S5.

| ***High Disembodiment Rest*** **From →To** | **Mean (Hz)** | **CI Low** | **CI High** | **Change (Hz)** | **CI Low** | **CI High** | **Mean + Change** |
|---|---|---|---|---|---|---|---|
| lAG → rTPJ | -0.181 | -0.251 | -0.111 | 0.189 | 0.092 | 0.285 | 0.008 |
| lAI → lAI | -0.235 | -0.354 | -0.116 | -0.261 | -0.433 | -0.089 | -0.496 |
| lAI → rAG | -0.12 | -0.193 | -0.047 | -0.16 | -0.266 | -0.053 | -0.28 |
| lTPJ → rAG | 0.109 | 0.041 | 0.178 | -0.119 | -0.216 | -0.021 | -0.01 |
| rAI → rAI | -0.168 | -0.298 | -0.038 | -0.349 | -0.531 | -0.167 | -0.517 |
| rAI → rTPJ | 0.125 | 0.031 | 0.218 | -0.173 | -0.304 | -0.043 | -0.048 |
| rTPJ → rAG | -0.124 | -0.19 | -0.058 | 0.184 | 0.094 | 0.273 | 0.06 |
| rTPJ → rAI | 0.12 | 0.053 | 0.186 | -0.115 | -0.211 | -0.02 | 0.005 |
| ***Music*** | | | | | | | |
| lAG → lAI | -0.138 | -0.209 | -0.067 | 0.196 | 0.096 | 0.296 | 0.058 |
| lAG → rAI | -0.074 | -0.131 | -0.017 | 0.146 | 0.064 | 0.229 | 0.072 |
| lTPJ → lAI | 0.098 | 0.017 | 0.18 | -0.158 | -0.281 | -0.035 | -0.06 |
| rTPJ → rAI | 0.279 | 0.207 | 0.35 | -0.133 | -0.238 | -0.029 | 0.146 |
| rTPJ → rTPJ | -0.154 | -0.26 | -0.047 | 0.245 | 0.092 | 0.399 | 0.091 |
| ***Low Disembodiment Rest*** | | | | | | | |
| lTPJ → lAG | -0.223 | -0.312 | -0.134 | 0.147 | 0.021 | 0.272 | -0.076 |
| lTPJ → lTPJ | -0.275 | -0.402 | -0.148 | -0.189 | -0.372 | -0.007 | -0.464 |
| lTPJ → rAG | -0.11 | -0.179 | -0.041 | 0.119 | 0.021 | 0.218 | 0.009 |
| rAG → rAG | -0.578 | -0.703 | -0.454 | 0.172 | 0 | 0.345 | -0.406 |
| rAI → rAG | -0.237 | -0.331 | -0.143 | 0.146 | 0.013 | 0.279 | -0.091 |
| rTPJ → rTPJ | -0.155 | -0.272 | -0.038 | 0.209 | 0.044 | 0.375 | 0.054 |
| ***Music*** | | | | | | | |
| lAI → rAG | -0.2 | -0.289 | -0.11 | -0.161 | -0.289 | -0.034 | -0.361 |
| rAI → lAI | 0.141 | 0.041 | 0.241 | -0.288 | -0.435 | -0.14 | -0.147 |

| | | | | | | | |
|---|---|---|---|---|---|---|---|
| rAI → rAG | -0.222 | -0.312 | -0.132 | -0.167 | -0.295 | -0.039 | -0.389 |

*Table S5. Mean effective connectivity (baseline) posterior probabilities displayed alongside change. lAG = left Angular Gyrus, rAG = right Angular Gyrus, lAI = left Anterior Insula, rAI = right Anterior Insula, lTPJ = left Temporoparietal Junction, and rTPJ = right Temporoparietal Junction. CI = confidence interval. Connections that produced a posterior probability .99 or greater in both mean effective connectivity and change are reported. Mean + Change estimates the mean effective connectivity under psilocybin. Underlined values indicate an estimated flip in the valence of the connection from mean (baseline) effective connectivity.*

*References*


1. Friston KJ, Kahan J, Biswal B, Razi A. A DCM for resting state fMRI. NeuroImage. 2014;94:396-407.
2. Beggs JM, Plenz D. Neuronal avalanches in neocortical circuits. J Neurosci. 2003;23(35):11167-77.
3. Shin CW, Kim S. Self-organized criticality and scale-free properties in emergent functional neural networks. Phys Rev E Stat Nonlin Soft Matter Phys. 2006;74(4 Pt 2):045101.
4. Stam CJ, de Bruin EA. Scale-free dynamics of global functional connectivity in the human brain. Hum Brain Mapp. 2004;22(2):97-109.
5. Razi A, Friston K. The Connected Brain: Causality, models, and intrinsic dynamics. IEEE Signal Processing Magazine. 2016;33(3):14-35.
6. Friston KJ, Harrison L, Penny W. Dynamic causal modelling. Neuroimage. 2003;19(4):1273-302.
7. Friston K, Mattout J, Trujillo-Barreto N, Ashburner J, Penny W. Variational free energy and the Laplace approximation. Neuroimage. 2007;34(1):220-34.
8. Friston KJ, Litvak V, Oswal A, Razi A, Stephan KE, van Wijk BC, et al. Bayesian model reduction and empirical Bayes for group (DCM) studies. Neuroimage. 2016;128:413-31.
9. Alexander Andrew S, Carstensen Lucas C, Hinman James R, Raudies F, Chapman GW, Hasselmo Michael E. Egocentric boundary vector tuning of the retrosplenial cortex. Science Advances. 2020;6(8):eaaz2322.
10. Kim M, Maguire EA. Hippocampus, retrosplenial and parahippocampal cortices encode multi-compartment 3D space in a hierarchical manner. bioRxiv. 2018:200576.
11. Vann SD, Aggleton JP, Maguire EA. What does the retrosplenial cortex do? Nature Reviews Neuroscience. 2009;10(11):792-802.
12. Doeller Christian F, Kaplan R. Parahippocampal Cortex: Translating Vision into Space. Current Biology. 2011;21(15):R589-R91.
13. Preller KH, Razi A, Zeidman P, Stämpfli P, Friston KJ, Vollenweider FX. Effective connectivity changes in LSD-induced altered states of consciousness in humans. Proceedings of the National Academy of Sciences. 2019;116(7):2743-8.
